# Causal associations between body fat accumulation and COVID-19 severity: A Mendelian randomization study

**DOI:** 10.1101/2022.01.20.22269593

**Authors:** Satoshi Yoshiji, Daisuke Tanaka, Hiroto Minamino, Takaaki Murakami, Yoshihito Fujita, J. Brent Richards, Nobuya Inagaki

## Abstract

**Purpose:** The causal effects of body fat mass and body fat-free mass on coronavirus disease 2019 (COVID-19) severity remain unclear. Here, we used Mendelian randomization (MR) to evaluate the causal relationships between body fat-related traits and COVID-19 severity.

**Material and Methods:** We identified single nucleotide polymorphisms associated with body mass index (BMI) and direct measures of body fat (i.e., body fat percentage, body fat mass, and body fat-free mass) in 461,460, 454,633, 454,137, and 454,850 individuals of European ancestry from the UK Biobank, respectively. We then performed two-sample MR to ascertain their effects on severe COVID-19 (cases: 4,792; controls: 1,054,664) from the COVID-19 Host Genetics Initiative.

**Results:** We found that an increase in BMI, body fat percentage, and body fat mass by one standard deviation were each associated with severe COVID-19 (odds ratio (OR)_BMI_ = 1.49, 95%CI: 1.19–1.87, *P* = 5.57×10^−4^; OR_body fat percentage_ = 1.94, 95%CI: 1.41–2.67, *P* = 5.07×10^−5^; and OR_body fat mass_ = 1.61, 95%CI: 1.28–2.04, *P* = 5.51×10^−5^). Further, we evaluated independent causal effects of body fat mass and body fat-free mass using multivariable MR and revealed that only body fat mass was independently associated with severe COVID-19 (OR_body fat mass_ = 2.91, 95%CI: 1.71–4.96, *P* = 8.85×10^−5^ and OR_body fat-free mass_ = 1.02, 95%CI: 0.61–1.67, *P* = 0.945).

**Conclusions:** This study demonstrates the causal effects of body fat accumulation on COVID-19 severity and indicates that the biological pathways influencing the relationship between COVID-19 and obesity are likely mediated through body fat mass.

## Introduction

Coronavirus disease-19 (COVID-19) has infected hundreds of millions of individuals and caused millions of deaths worldwide (1). The severity of COVID-19 varies considerably among individuals, and identifying modifiable risk factors associated with COVID-19 severity is essential for optimizing public health policies, allocating resources, and assisting clinical decisions.

A major risk factor for COVID-19 appears to be obesity. A community-based cohort study involving 6.9 million individuals in England showed a positive association between body mass index (BMI) and COVID-19 severity (2). In that study, BMI was significantly associated with hospital admission, admission to an intensive care unit, and death due to COVID-19, with these findings being replicated in other independent observational studies (3, 4). However, these results do not support causation; in fact, interpreting these observations as a causal relationship relies on untestable and usually implausible assumptions, including the absence of unmeasured confounders and reverse causation (5). Given these limitations inherent to traditional observational epidemiology studies, Mendelian randomization (MR) has emerged as a way to mitigate against such shortcomings through its use of genetic variants as instrumental variables in order to infer a causal relationship between exposures and outcomes (6, 7). Using MR, we can estimate the causal effects of genetically predicted levels of adiposity-related exposures on COVID-19 outcomes, in contrast to typical observational studies that evaluate only associations. Because genetic alleles are randomly assigned at conception, which is generally well before the onset of the disease, the risk of reverse causation is substantially decreased.

The selection of proxy measures of adiposity plays a vital role in evaluating the association between obesity and COVID-19 outcomes. BMI can be measured easily and is therefore a common measurement of obesity in epidemiological studies. However, the key limitation of BMI is that it is an indirect measure of obesity because it is calculated only with height and weight and does not consider body composition (i.e., body fat mass, body fat-free mass, and their ratio) (8). Therefore, direct measures of body fat accumulation (i.e., body fat percentage and body fat mass) might better elucidate the association of body fat with COVID-19 outcomes. Taking this into consideration, we used a dataset of the UK Biobank, which is a prospective cohort in the UK involving ∼500,000 individuals (463,844 of which are of European ancestry) with detailed genetic and phenotypic information. Data from the UK Biobank includes measurements of not only BMI but also body fat mass and body fat-free mass acquired through the bioelectrical impedance analysis and dual-energy X-ray absorptiometry along with the results from the genome-wide association studies (GWAS) for these traits (9). Recent MR studies utilized the results from the UK Biobank to obtain single nucleotide polymorphisms (SNPs) associated with body fat accumulation and showed estimated causal effects of body fat on various traits ranging from cardiovascular diseases to depression (10-13). In light of these promising findings, body fat mass has emerged as a valuable indicator of the deleterious effects of fat accumulation; however, it remains unclear whether fat mass and fat percentage are causally related to severe outcomes of COVID-19.

In this study, we therefore conducted a two-sample MR to assess whether BMI and direct measures of body compositions (i.e., body fat percentage, body fat mass, and body fat-free mass) are causally associated with severe COVID-19 and COVID-19 hospitalization using data from the UK Biobank and the COVID-19 Host Genetics Initiative.

## Methods

### Instrumental variables for BMI, body fat percentage, body fat mass, and body fat-free mass

Instrumental variables were defined as independent genome-wide significant SNPs (*P* < 5×10^−8^) for exposure traits. Independence of SNPs was defined as not in linkage disequilibrium with other SNPs (*r*^*2*^ < 0.001 within a 10,000 kb window). The exposures used in this study were BMI, body fat percentage, body fat mass and body fat-free mass. To select SNPs used as instrumental variables, we obtained the GWAS results of BMI, body fat percentage, body fat mass, and body fat-free mass from individuals with European ancestry in the UK Biobank (**Figure 1**), using the OpenGWAS and MR-Base platform of the MRC Integrative Epidemiology Unit at the University of Bristol (14). The fat mass and fat-free mass of the UK Biobank participants were evaluated by performing bioelectrical impedance analysis using the Tanita BC418MA body composition analyzer (Tanita, Tokyo, Japan). We restricted the analyses to individuals of European ancestry in order to maximize the statistical power, given that the majority of UK Biobank participants were of European ancestry. To select instrumental variables, SNPs were clumped using PLINK (v1.90) according to a linkage disequilibrium threshold of *r*^*2*^ < 0.001 with a clumping window of 10,000 kb using the 1000G European reference panel (9, 14) in order to select an independent SNP with the lowest *P*-value in each linkage disequilibrium block. When a selected SNP was not present in the results of the GWAS on COVID-19 severity outcomes, we instead used a proxy SNP that was in linkage disequilibrium with the selected SNP instead, with an *r*^*2*^ of ≥0.8 and minor allele frequency of ≤0.3 using 1000G European reference panel as described elsewhere (12). We calculated *F*-statistics for the exposure traits and a genetic correlation between body fat mass and body fat-free mass using LDAK (v5.1) (15).

**Figure 1.**
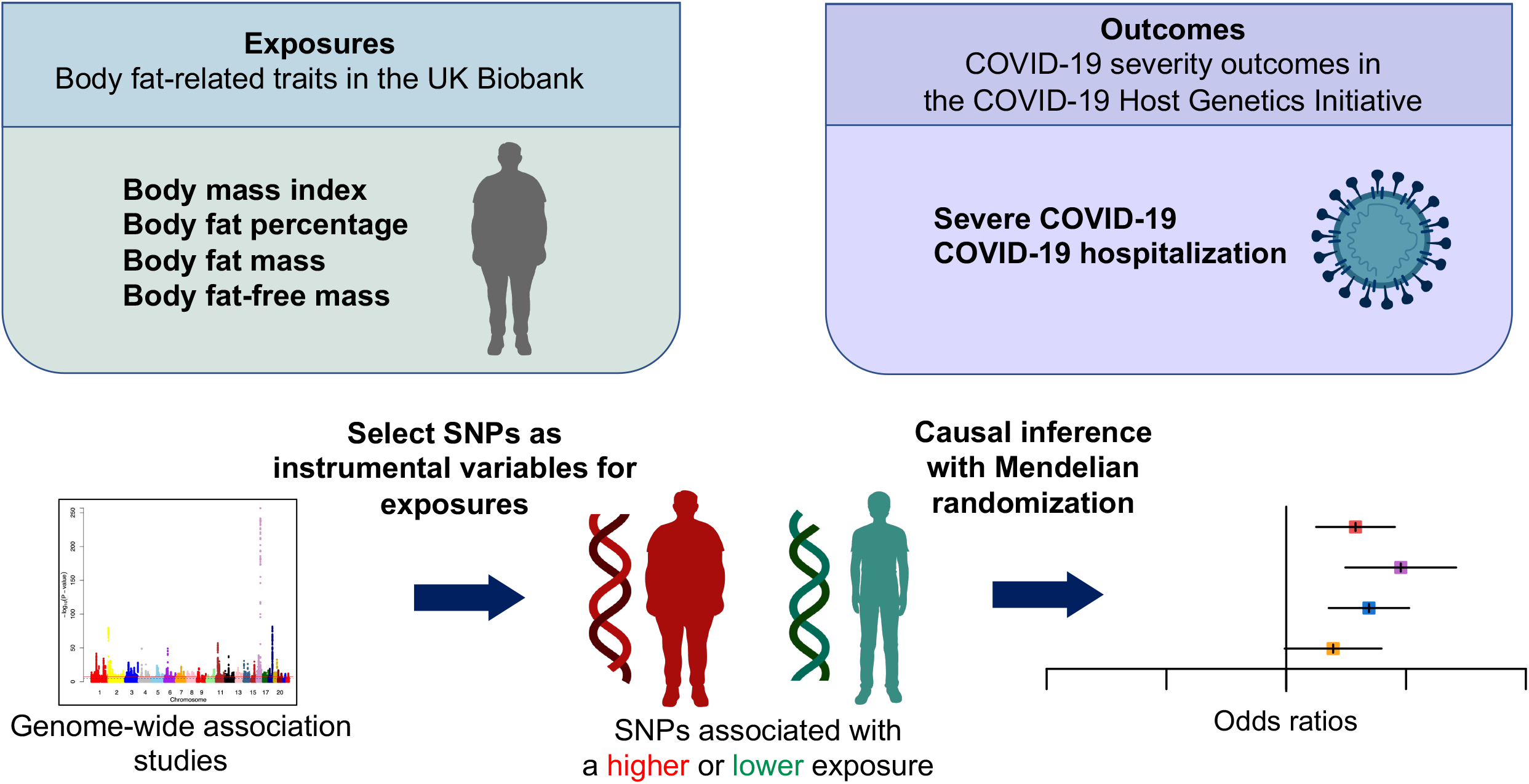
Schematic representation of the Mendelian randomization study. SNPs = single nucleotide polymorphisms.

### Severe COVID-19 and COVID-19 hospitalization outcomes

For proxy outcomes of COVID-19 severity, we adopted the outcomes of the COVID-19 Host Genetics Initiative, which is an international consortium that works collaboratively to share data and ideas, recruit patients, and disseminate scientific findings. The outcomes were severe COVID-19 and COVID-19 hospitalization (16). For definitions of COVID-19 outcomes, the severe COVID-19 group was defined as individuals whose death was due to COVID-19, those requiring respiratory support, or those requiring hospitalization due to symptoms related to laboratory-confirmed SARS-CoV-2 infection. The COVID-19 hospitalization group was defined as individuals requiring hospitalization due to symptoms associated with laboratory-confirmed SARS-CoV-2 infection. We used the largest GWAS summary statistics of the COVID-19 Host Genetics Initiative for severe COVID-19 and COVID-19 hospitalization outcomes in individuals of European-ancestry, excluding those from the UK Biobank. We restricted the population to those of European ancestry in order to minimize confounding due to population stratification. The datasets corresponding to each outcome were as follows: severe COVID-19 (cases: 4,792; controls: 1,054,664; dataset ID: COVID19_HGI_A2_ALL_eur_leave_ukbb_23andme_20210107) and COVID-19 hospitalization (cases: 14,652; controls: 1,114,836; and dataset ID: COVID19_HGI_B2_ALL_eur_leave_ukbb_23andme_20210622). For the definitions of controls in the GWAS data, ancestry-matched controls were sourced from participating population-based cohorts. Controls included individuals whose status of exposure to SARS-CoV-2 was either negative according to electronic health records/questionnaires or unknown (16).

### Mendelian randomization

We performed univariable MR using the inverse-variance weighted method (hereinafter referred to as univariable MR) to evaluate the relationship of BMI, body fat percentage, body fat mass, and body fat-free mass with severe COVID-19 and COVID-19 hospitalization. Univariable MR is a weighted linear regression model in which the effect of genetic variants *i* (*i*= 1 … *n*) on an outcome 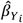 is regressed on the effect of the same genetic variant *i* on the exposure 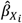 weighted by the inverse of the squared standard error 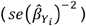. The estimated total effect (*θ*) of the exposure on the outcome can be formulated as follows:

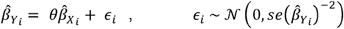

The instrumental variable assumptions are: (I) Relevance: genetic variant is associated with the exposure. (II) Independence: genetic variant does not share the unmeasured cause or confounder with the outcome. (III) Exclusion restriction: genetic variant does not influence the outcome except through the exposure (6, 7). These assumptions are illustrated by a canonical diagram in **Figure 2**.

**Figure 2.**
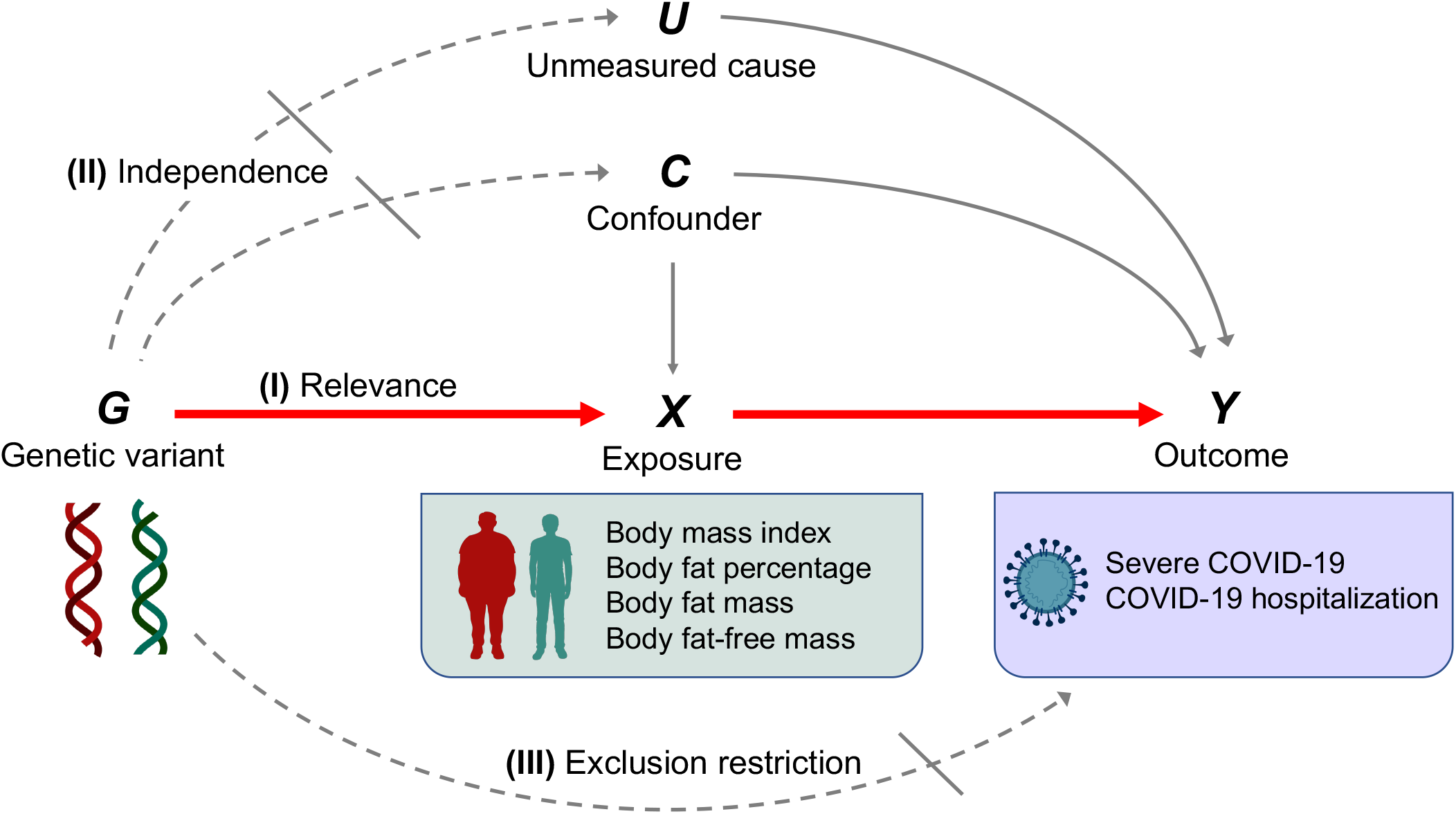
Canonical diagram illustrating the instrumental variable assumptions made in the Mendelian randomization analyses. Genetic variant ***G*** is used as an instrumental variable for exposure ***X*** (body mass index, body fat percentage, body fat mass, or body fat-free mass) to evaluate the causal effect of ***X*** on the outcome ***Y*** (severe COVID-19 or COVID-19 hospitalization). Instrumental variable assumptions include: (I) Relevance: genetic variant ***G*** is associated with exposure ***X***. (II) Independence: genetic variant ***G*** does not share the unmeasured cause or the confounder with the outcome ***Y***. (III) Exclusion restriction: genetic variant ***G*** does not influence the outcome ***Y*** except through the exposure ***X***. Red solid arrows represent causal effects; grey solid arrows represent causal effects of the unmeasured cause or confounder that do not violate the instrumental variable assumptions; dashed arrows represent causal effects that are specifically prohibited by the instrumental variable assumptions.

Multivariable MR was performed using the inverse-variance weighted method (hereinafter referred to as multivariable MR). This is an extension of univariable MR, in which the effects of genetic variant *i* (*i*= 1 … *n*) on the outcome 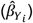 are regressed on the effect of genetic variant *i* on two exposures of *X*_*1*_ (fat mass) and *X*_*2*_ (fat-free mass). In multivariable MR, genetic variants used as instrumental variables are associated with one or both of the exposures (17).

The causal associations were evaluated using odds ratios (ORs), which are expressed according to a standard deviation (SD) increase in genetically predicted BMI (kg/m^2^), body fat percentage (%), body fat mass (kg), or body fat-free mass (kg).

Results with a *P* < 0.0125 were considered statistically significant (*P* = 0.05/4; Bonferroni-corrected significance threshold according to the number of exposures). We note that such a correction is likely overly conservative, given that the exposures are non-independent. MR analyses were performed using TwoSampleMR (v0.5.6) in R (v4.02). This study is conducted in accordance with the STROBE-MR guideline (6, 7). STROBE-MR checklist is provided in **Supplementary material** (18).

### Sensitivity analysis

We performed the MR-Egger intercept test, Cochran’s Q test, and the MR-PRESSO global test (19, 20) to detect horizontal pleiotropy, which occurs when instrumental variables influence outcomes through pathways independent of the exposure. MR-Egger relaxes the exclusion restriction assumption and is valid under the Instrument Strength Independent of Direct Effect (InSIDE) assumption that associations of the genetic variants with the exposure trait are independent of direct effects of the genetic variants on the outcome. Deviation of the MR-Egger intercept from zero indicates horizontal pleiotropy. The results of Cochran’s Q test were used to evaluate the heterogeneity of genetic variants used as instrumental variables. Results of Cochran’s Q test were presented with *I*^*2*^ index, based on which the heterogeneity of genetic variants was defined categorically with *I*^*2*^ index as low (*I*^*2*^ index ≤ 25%), moderate (*I*^*2*^ index 26–50%), and high (*I*^*2*^ index > 50%). Additionally, we performed the MR-PRESSO global test, which can detect horizontally pleiotropic outlier SNPs. A significant result indicates the presence of pleiotropic outlier SNPs and this method then generates ORs after removing and correcting for these outliers (outlier-corrected ORs). MR-PRESSO can also be used to evaluate the distortion of the causal estimates before and after the removal of pleiotropic outlier SNPs following the MR-PRESSO distortion test. MR-PRESSO requires at least 50% of the genetic variants to be valid instruments with no horizontal pleiotropy and also relies on the InSIDE assumption.

Results with a *P* < 0.05 were considered to indicate the presence of horizontal pleiotropy for the MR-Egger intercept test, Cochran’s Q test, MR-PRESSO global test, and MR-PRESSO distortion test. Sensitivity analyses were performed with TwoSampleMR (v.0.5.6) and MR-PRESSO (v1.0).

### Ethics statements

The UK Biobank and COVID-19 Host Genetics Initiatives obtained ethics approval from the relevant institutional ethics committees. We used publicly available summary statistics of GWAS results of UK Biobank and COVID-19 Host Genetics Initiative and did not use individual-level data.

## Results

### Instrumental variables for exposure traits

The characteristics of the exposure traits (BMI, body fat percentage, body fat mass, and body fat-free mass) are presented in **Table 1**. The mean ± SD BMI was 27.4 ± 4.8 kg/m^2^, body fat percentage was 31.4 ± 8.5%, body fat mass was 24.9 ± 9.6 kg, and body fat-free mass was 53.2 ± 11.5 kg (**Table 1**). For BMI, body fat percentage, body fat mass, and body fat-free mass, 439, 377, 417, and 530 independent genome-wide significant SNPs were identified as instrumental variables from the GWAS results of the UK Biobank, respectively. *F*-statistics for these exposure traits were 507.6, 496.9, 502.2, and 607.4, respectively. The SNPs used as instrumental variables are presented in **Supplementary Table 1** (18).

**Table 1.** Dataset descriptions.

| Data source | Dataset details | Phenotype | Sample size of each dataset | Mean ± SD |
| --- | --- | --- | --- | --- |
| UK Biobank | • GWAS in individuals of European ancestry.<br>• Body fat and body fat-free mass were measured using bioelectrical impedance analysis. | Body mass index | 461,460 | 27.4 ± 4.8 kg/m<sup>2</sup> |
| | | Body fat percentage | 454,633 | 31.4 ± 8.5% |
| | | Body fat mass | 454,137 | 24.9 ± 9.6 kg |
| | | Body fat-free mass | 454,850 | 53.2 ± 11.5 kg |
| COVID-19 Host Genetics Initiative | • Meta-analysis of GWAS in individuals of European ancestry excluding those from UK biobank | Severe COVID-19 | Cases: 4,792<br>Controls: 1,054,664 | – |
|  |  | COVID-19 hospitalization | Cases: 14,652<br>Controls: 1,114,836 | – |

### Severe COVID-19 outcome

For the severe COVID-19 outcome (**Figure 3**), univariable MR showed that the genetically predicted increase per SD in BMI, body fat percentage, and body fat mass was associated with an increased risk of severe COVID-19 (OR_BMI_ = 1.49, 95%CI: 1.19–1.87, *P* = 5.57×10^−4^; OR_fat percentage_ = 1.94, 95%CI: 1.41–2.67; *P* = 5.07×10^−5^, OR_body fat mass_ = 1.61, 95%CI 1.28–2.04, *P* = 5.51×10^−5^; and OR_body fat-free mass_ =1.31, 95%CI: 0.99–1.74, *P* = 5.77×10^−2^). Further, as instrumental variables for body fat mass and body fat-free mass were not independent from each other (*r*^*2*^ *=* 0.64 for the genetic correlation of the two traits), we performed multivariable MR to elucidate the independent causal effects of body fat mass and body fat-free mass on the severe COVID-19 outcome, which showed that only body fat mass was independently associated with the severe COVID 19 outcome (body fat mass: OR_body fat mass_ = 2.91, 95%CI: 1.71–4.96, *P* = 8.85×10^−5^, and OR_body fat-free mass_ = 1.02, 95%CI: 0.61–1.67, *P* = 0.945).

**Figure 3.**
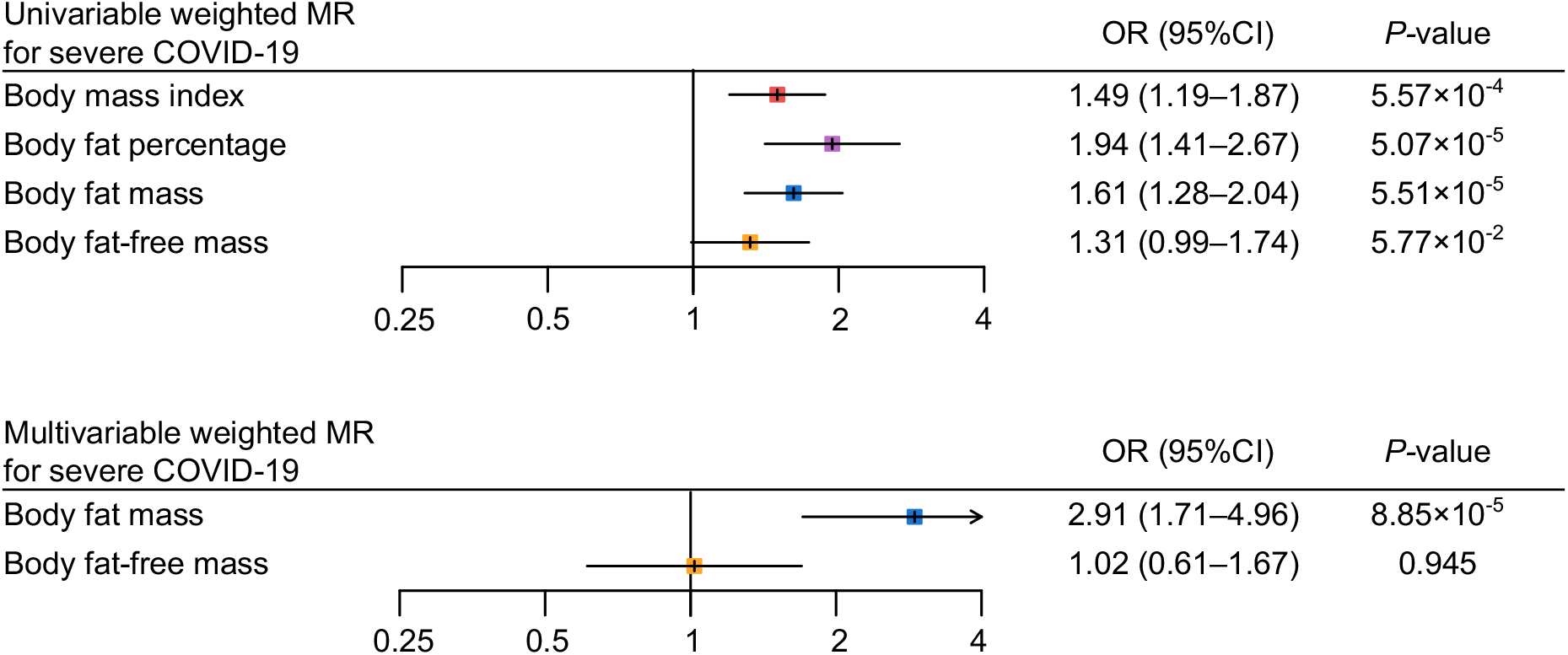
Univariable and multivariable Mendelian randomization analysis of the severe COVID-19 outcome. MR = Mendelian randomization.

### COVID-19 hospitalization outcome

For the COVID-19 hospitalization outcome (**Figure 4**), univariable MR showed that a genetically predicted increase per SD in BMI, body fat percentage, body fat mass, and body fat-free mass was associated with an increased risk of COVID-19 hospitalization (OR_BMI_ = 1.31, 95%CI: 1.19–1.44, *P* = 3.46 × 10^−8^; OR_fat percentage_ = 1.44, 95%CI: 1.26–1.66, *P* = 1.22 × 10^−7^; OR_body fat mass_ = 1.32, 95%CI: 1.20–1.46, *P* = 2.52 × 10^−8^; OR_body fat-free mass_ = 1.27 95%CI: 1.13–1.42, *P* = 4.44 × 10^−5^). In multivariable MR, only body fat mass was independently associated with COVID-19 hospitalization (OR_body fat mass_ = 2.38, 95%CI: 1.56–3.61, *P* = 5.29×10^−5^; OR_body fat-free mass_ = 0.82, 95%CI: 0.56–1.19, *P* = 0.293), consistent with the findings for severe COVID-19.

**Figure 4.**
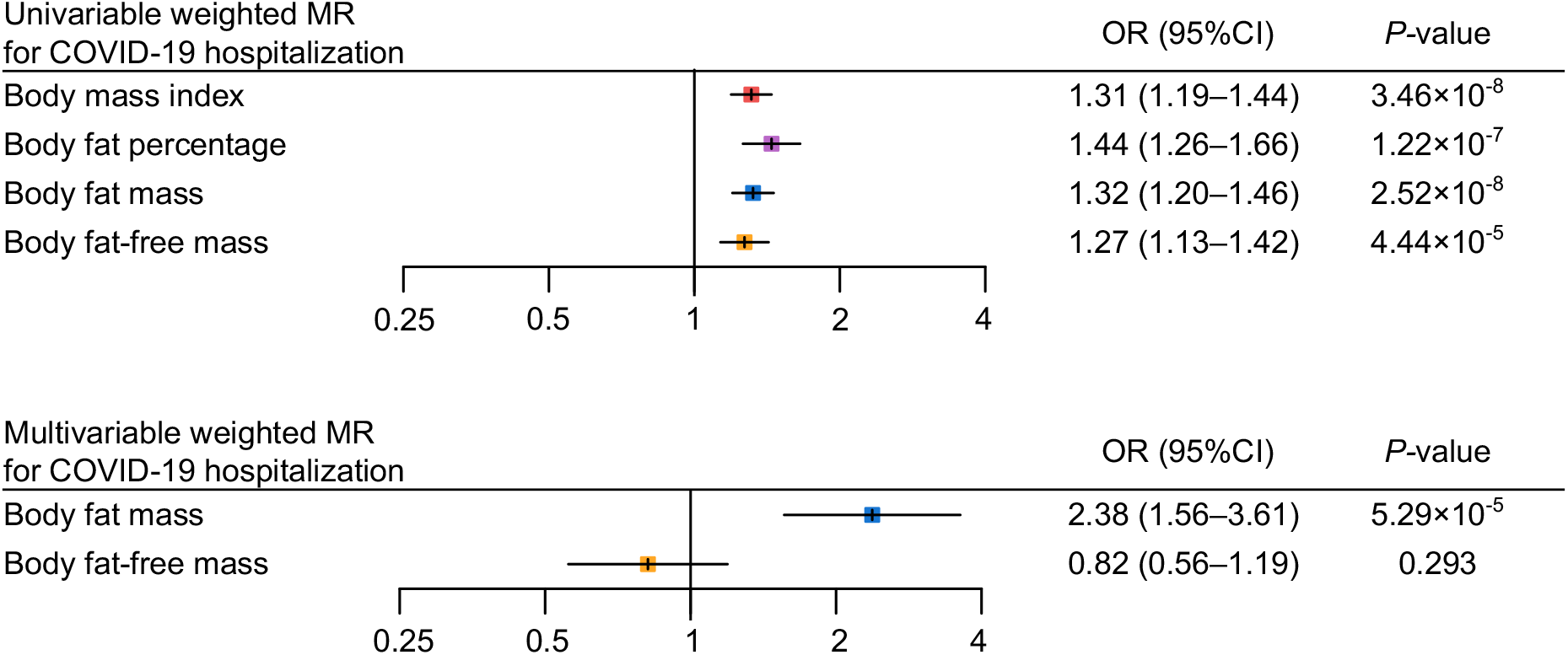
Univariable and multivariable Mendelian randomization analysis for the COVID-19 hospitalization outcome. MR = Mendelian randomization.

### Sensitivity analysis

We performed MR-Egger, Cochran’s Q test and MR-PRESSO for sensitivity analysis (**Table 2**). In the MR-Egger, the 95%CI results of the MR-Egger intercept (Egger-intercept) contained the null hypothesis value zero for all exposure-outcome relationships, suggesting no evidence of horizontal pleiotropy. Heterogeneity estimates of instrumental variables were low according to the *I*^*2*^ index (*I*^*2*^ index were ≤ 25% for all exposure traits). However, MR-PRESSO detected some pleiotropic outlier SNPs in instrumental variables for BMI, body fat percentage, and body fat mass with the COVID-19 hospitalization outcome (*P*-value for global test < 0.05). Nevertheless, results with MR-PRESSO after removal and correction for these pleiotropic outlier SNPs were directionally consistent with those from univariable MR, supporting the robustness of the findings with univariable MR. In addition, the MR-PRESSO distortion test detected no significant distortion in the causal estimates before and after removal of outlier pleiotropic SNPs.

**Table 2.**
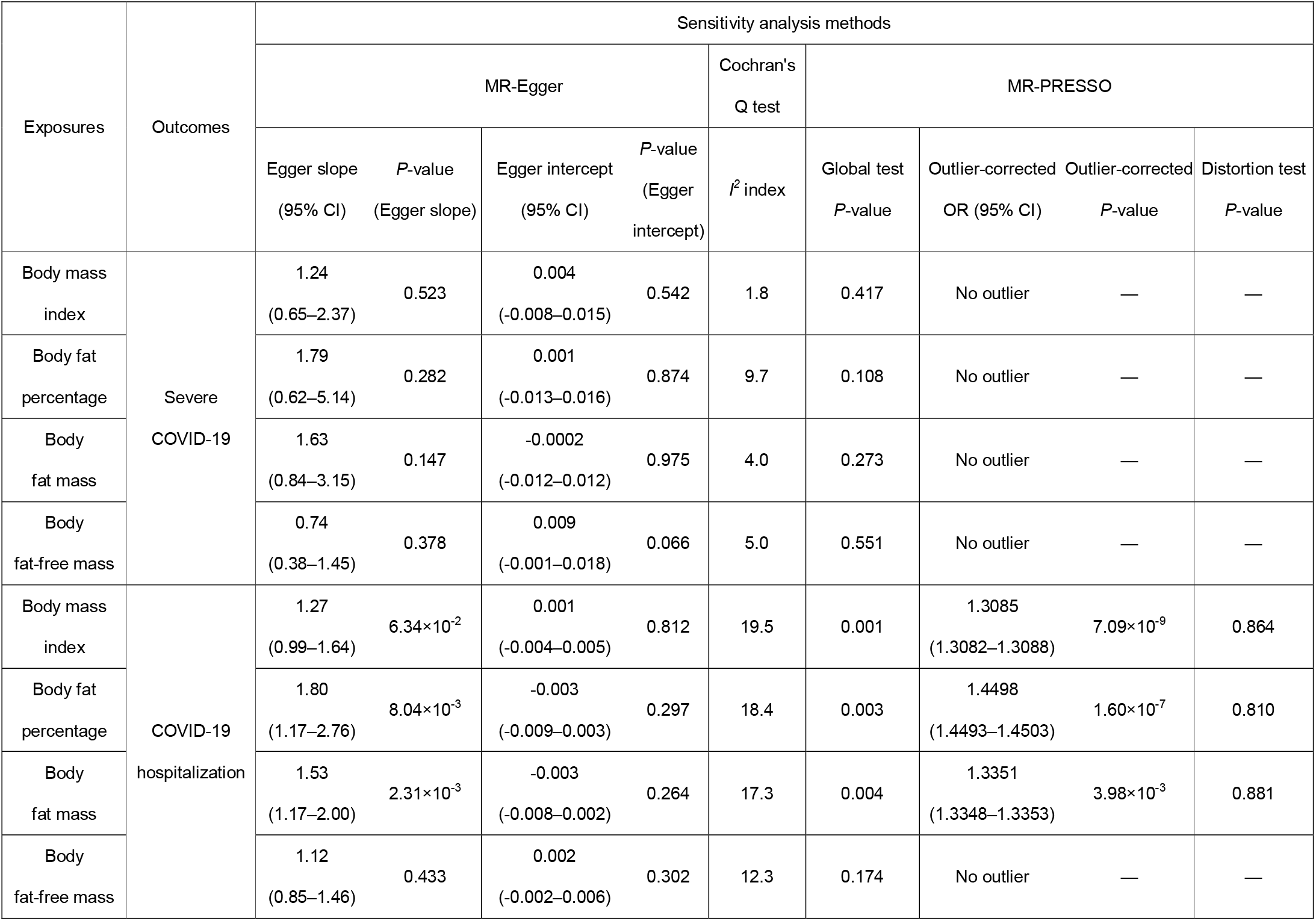
Sensitivity analysis results.

| Exposures | Outcomes | Sensitivity analysis methods |  |  |  |  |  |  |  |  |
| --- | --- | --- | --- | --- | --- | --- | --- | --- | --- | --- |
|  |  | MR-Egger |  |  |  | Cochran's<br>Q test | MR-PRESSO |  |  |  |
|  |  | Egger slope<br>(95% CI) | *P*-value<br>(Egger slope) | Egger intercept<br>(95% CI) | *P*-value<br>(Egger<br>intercept) | *I*<sup>2</sup> index | Global test<br>*P*-value | Outlier-corrected<br>OR (95% CI) | Outlier-corrected<br>*P*-value | Distortion test<br>*P*-value |
| Body mass<br>index | Severe<br>COVID-19 | 1.24<br>(0.65–2.37) | 0.523 | 0.004<br>(-0.008–0.015) | 0.542 | 1.8 | 0.417 | No outlier | — | — |
| Body fat<br>percentage |  | 1.79<br>(0.62–5.14) | 0.282 | 0.001<br>(-0.013–0.016) | 0.874 | 9.7 | 0.108 | No outlier | — | — |
| Body<br>fat mass |  | 1.63<br>(0.84–3.15) | 0.147 | -0.0002<br>(-0.012–0.012) | 0.975 | 4.0 | 0.273 | No outlier | — | — |
| Body<br>fat-free mass |  | 0.74<br>(0.38–1.45) | 0.378 | 0.009<br>(-0.001–0.018) | 0.066 | 5.0 | 0.551 | No outlier | — | — |
| Body mass<br>index | COVID-19<br>hospitalization | 1.27<br>(0.99–1.64) | 6.34×10<sup>-2</sup> | 0.001<br>(-0.004–0.005) | 0.812 | 19.5 | 0.001 | 1.3085<br>(1.3082–1.3088) | 7.09×10<sup>-9</sup> | 0.864 |
| Body fat<br>percentage |  | 1.80<br>(1.17–2.76) | 8.04×10<sup>-3</sup> | -0.003<br>(-0.009–0.003) | 0.297 | 18.4 | 0.003 | 1.4498<br>(1.4493–1.4503) | 1.60×10<sup>-7</sup> | 0.810 |
| Body<br>fat mass |  | 1.53<br>(1.17–2.00) | 2.31×10<sup>-3</sup> | -0.003<br>(-0.008–0.002) | 0.264 | 17.3 | 0.004 | 1.3351<br>(1.3348–1.3353) | 3.98×10<sup>-3</sup> | 0.881 |
| Body<br>fat-free mass |  | 1.12<br>(0.85–1.46) | 0.433 | 0.002<br>(-0.002–0.006) | 0.302 | 12.3 | 0.174 | No outlier | — | — |

## Discussion

To the best of our knowledge, this is the first MR study to evaluate the causal association between directly measured body compositions (i.e., body fat percentage, body fat mass, and body fat-free mass) with COVID-19 severity outcomes. In this study, we found that an increase in BMI, body fat percentage, and body fat mass were associated with an increased risk of severe COVID-19 and COVID-19 hospitalization. We further evaluated the independent causal effects of body fat mass and body fat-free mass on these outcomes and revealed that only body fat mass was independently associated with the outcomes.

During the COVID-19 pandemic, obesity has emerged as a major risk factor for COVID-19 outcomes. Multiple observational studies showed that obese individuals present an increased risk of severe diseases, hospitalization, and death due to COVID-19 (2, 3, 21, 22). However, observational studies are prone to confounding bias and reverse causation and do not estimate the causal effects of exposures on outcomes. To overcome these limitations, several MR studies have been performed to evaluate the causal relationship between obesity-related traits and COVID-19 outcomes. Among anthropometric traits including BMI, waist circumference, hip circumference, and waist-to-hip ratio, BMI showed an association with poor COVID-19 outcomes (23). However, BMI is calculated only from height and weight and does not consider body compositions. Moreover, these anthropometric traits are indirect measures of obesity and might not be accurate proxies for body fat. Therefore, it is necessary to evaluate associations between directly measured fat traits (i.e., body fat mass and body fat percentage) and COVID-19 severity. In this MR study, we adopted these directly measured traits as exposure traits and provided novel findings that indicate the causal association of body fat accumulation with severe COVID-19 outcome.

We used multivariable MR since most instrumental variables of adiposity effect both fat mass and fat-free mass, although some variants more strongly and proportionally influence fat mass, whereas others influence fat-free mass more strongly. Therefore, multivariable MR can test the differential causal effects of fat mass and fat-free mass. Using this approach, recent MR studies showed differential associations between body fat mass and body fat-free mass with various disorders (10-13). The present findings extend this knowledge to COVID-19. Results from multivariable MR showed that body fat mass but not body fat-free mass was independently associated with severe COVID-19 and COVID-19 hospitalization. The association between body fat mass and COVID-19 severity was strengthened in multivariable MR relative to findings using univariable MR, whereas the effects of body fat-free mass on COVID-19 severity was markedly attenuated in multivariable MR, thereby illustrating the independent associations between body fat mass and COVID-19 severity.

The underlying mechanism of these associations remains to be clarified. Obesity is a metabolic disease characterized by systemic changes in metabolism, including insulin resistance, glucose intolerance, dyslipidemia, changes in adipokines (e.g., increased leptin and decreased adiponectin levels), chronic inflammation, and altered immune response, all of which could collectively increase the risk of COVID-19 severity (4, 24, 25). Moreover, obesity causes respiratory dysfunction, including impaired respiratory physiology, increased airway resistance, impaired gas exchange, low lung volume, and low muscle strength, which can also increase the risk of COVID-19 severity. Furthermore, the physical characteristics of obese individuals render intubation and laryngoscopy difficult, which could also aggravate outcomes (26). Further studies are needed to explore the pathways linking adiposity to increased risk of COVID-19 severity.

This study has several strengths. We used an MR design, which minimized bias from reverse causation and confounders, thereby enabling us to test for causal effects, provided compliance with MR assumptions. In this MR study, we used the data from the UK Biobank for the exposure traits (*F*-statistics > 10 for all exposure traits) and COVID-19 Host Genetics Initiative for the outcomes, both of which have large sample sizes, thus increasing the statistical power of the analysis. Furthermore, as proxy measures of body compositions, we not only considered BMI, which is a common indirect measure, but also direct measures, including body fat percentage, body fat mass, and body fat-free mass, and revealed associations of these traits with COVD-19 severity.

However, our study also has important limitations. First, the MR analysis relies on several key assumptions, the violation of which compromises causal inference. To test for possible violations of these assumptions, we performed multiple sensitivity analyses. The MR-Egger intercept test did not detect horizontal pleiotropy. Although SNP heterogeneities were detected when analyzing the outcome in terms of COVID-19 hospitalizations, the removal of outlier SNPs via MR-PRESSO still showed results consistent with those from MR with the inverse-variance method. We believe that these sensitivity analyses demonstrate the robustness and validity of the present findings, however we acknowledge that horizontal pleiotropy is difficult to exclude entirely. Second, regarding exposure traits, we used measures derived from the bioelectrical impedance analysis (i.e., body fat percentage, body fat mass, and body fat-free mass) instead of dual-energy X-ray absorptiometry (DXA)-derived measures to maximize statistical power. Although the UK Biobank collected DXA-derived measures for body fat mass, and body fat-free mass, the sample size was markedly smaller for these measurements (*n* = 5,170). Moreover, although DXA-derived measures are generally more accurate than impedance-derived measures, high correlations between the two were reported for fat mass (*r* = 0.96) and fat-free mass (*r* = 0.86) in the UK Biobank dataset (10). Hence, we believe impedance-derived measures can serve as clinically-relevant exposure traits in the present analysis. Third, we only used summary-level data and did not use individual-level data. Therefore, we could not evaluate the nonlinear relationship between exposures and outcomes. However, it should be noted that MR using summary statistics can still test for the presence of causal effects of exposures on outcomes, even if the exposure-outcome relationship is nonlinear (27). Additionally, a recent prospective cohort study of 6.9 million individuals in the UK suggested that BMI and COVID-19 severity show a linear relationship within a BMI range ≥23 kg/m^2^ (2). Notably, the BMI of a majority of the individuals in the UK Biobank population included in the present analysis fell within this range (≥23 kg/m^2^). Fourth, we restricted our analysis to individuals of European ancestry given that majority of participants in the UK Biobank were of European ancestry. Future studies are warranted to evaluate the generalizability of our findings to other populations.

In summary, the present MR study provides evidence that indicates a causal relationship between body fat accumulation and COVID-19 severity. Because excess fat can be reduced by following an appropriate diet and exercising, it might represent an important modifiable risk factor. Thus, body weight reduction considering direct measurements of body fat (i.e., body fat percentage and body fat mass) can be an effective strategy to reduce the risk of COVID-19 severity.

## Supporting information

https://figshare.com/s/c876361a354038ea988f

## Data Availability

All GWAS summary statistics used in this study are publicly available. All results are included in the present article.

## Acknowledgments

The Richards research group is supported by the Canadian Institutes of Health Research (CIHR: 365825; 409511, 100558, 169303), the McGill Interdisciplinary Initiative in Infection and Immunity (MI4), the Lady Davis Institute of the Jewish General Hospital, the Jewish General Hospital Foundation, the Canadian Foundation for Innovation, the NIH Foundation, Cancer Research UK, Genome Québec, the Public Health Agency of Canada, McGill University, Cancer Research UK [grant umber C18281/A29019] and the Fonds de Recherche Québec Santé (FRQS). JBR is supported by a FRQS Mérite Clinical Research Scholarship. Support from Calcul Québec and Compute Canada is acknowledged. TwinsUK is funded by the Welcome Trust, Medical Research Council, European Union, the National Institute for Health Research (NIHR)-funded BioResource, Clinical Research Facility and Biomedical Research Centre based at Guy’s and St Thomas’ NHS Foundation Trust in partnership with King’s College London. These funding agencies had no role in the design, implementation or interpretation of this study. SY and HM are supported by the Japan Society for the Promotion of the Science.

