## Supplementary material for "Causal associations between body fat accumulation and COVID-19 severity: A Mendelian randomization study": https://figshare.com/s/c876361a354038ea988f: STROBE-MR_Checklist.docx

| **Item** | **Page No. and Relevant text from manuscript** |
| --- | --- |
| **1. Title and Abstract:** Indicate Mendelian randomization (MR) as the study’s design in the title and/or the abstract if that is a main purpose of the study. | Page 1. Specified in the title and in the abstract. |
| **Introduction** |  |
| 2. **Background:** Explain the scientific background and rationale for the reported study. What is the exposure? Is a potential causal relationship between exposure and outcome plausible? Justify why MR is a helpful method to address the study question | Page 6–8. Explained in the 1^st^–3^rd^ paragraphs of the introduction. |
| 3. **Objectives:** State specific objectives clearly, including prespecified causal hypotheses (if any). State that MR is a method that, under specific assumptions, intends to estimate causal effects | Page 8. Stated in the 4^th^ paragraph of the introduction. |
| **Methods** |  |
| 4. **Study design and data sources:** Present key elements of the study design early in the article. Consider including a table listing sources of data for all phases of the study. For each data source contributing to the analysis, describe the following:   a) Setting: Describe the setting, locations, and relevant dates, including periods of recruitment, exposure, follow-up, and data collection, when available  b) Participants: Give the eligibility criteria, and the sources and methods of selection of participants. Report the sample size, and whether any power or sample size calculations were carried out prior to the main analysis  c) Describe measurement, quality control and selection of genetic variants  d) For each exposure, outcome, and other relevant variables, describe methods of assessment and diagnostic criteria for diseases  e) Provide details of ethics committee approval and participant informed consent, if relevant | Page 8–13. Presented exposures and outcomes in the “Instrumental variables for BMI, body fat percentage, body fat mass, and body fat-free mass“ section and the “Severe COVID-19 and COVID-19 hospitalization outcomes” section.  Also mentioned key design of the study in the “Mendelian randomization” section.  Included Figure 1 for describing the study design. Also included Table 1 for the dataset description. Elaborated on a)–e) in the “Instrumental variables for BMI, body fat percentage, body fat mass, and body fat-free mass” section, the “Severe COVID-19 and COVID-19 hospitalization outcomes” section, the “Mendelian randomization” section, and the “Ethical statements” section. |
| 5. **Assumptions:** Explicitly state the three core IV assumptions for the main analysis (relevance, independence and exclusion restriction) as well assumptions for any additional or sensitivity analysis | Page 10–13. Stated in the “Mendelian randomization” section and in Figure 2. |
| 6. **Statistical methods main analysis**   Describe statistical methods and statistics used  a) Describe how quantitative variables were handled in the analyses (that is, scale, units, model)  b) Describe how genetic variants were handled in the analyses and, if applicable, how their weights were selected  c) Describe the MR estimator (eg, two stage least squares, Wald ratio) and related statistics. Detail the included covariates and, in the case of two sample MR, whether the same covariate set was used for adjustment in the two samples  d) Explain how missing data were addressed.  e) If applicable, indicate how multiple testing was addressed | Page 10–13. Described in the "Mendelian randomization” section. |
| 7. **Assessment of assumptions:** Describe any methods used to assess the assumptions or justify their validity | Page 12–13. Described in the “Sensitivity analysis” section. |
| 8. **Sensitivity analyses:** Describe any sensitivity analyses or additional analyses performed (eg, comparison of effect estimates from different approaches, independent replication, bias analytic techniques, validation of instruments, simulations) | Page 12–13. Described in the “Sensitivity analysis” section. |
| 9. **Software and pre-registration**   a) Name statistical software and package(s), including version and settings used  b) State whether the study protocol and details were pre-registered (as well as when and where) | Page 8–13. Described in the “Instrumental variables for BMI, body fat percentage, body fat mass, and body fat-free mass” section and the “Mendelian randomization” section. |
| **Results** |  |
| 10. **Descriptive data**   a) Report the numbers of individuals at each stage of included studies and reasons for exclusion. Consider use of a flow-diagram  b) Report summary statistics for phenotypic exposure(s), outcome(s) and other relevant variables (e.g. means, standard deviations, proportions)  c) If the data sources include meta-analyses of previous studies, provide the number of studies, their reported ancestry, if available, and assessments of heterogeneity across these studies. Consider using a supplementary table for each data source  d) For two-sample Mendelian randomization:  i. Provide information on the similarity of the genetic variant-exposure associations between the exposure and outcome samples  ii. Provide information on extent of sample overlap between the exposure and outcome data sources | Page 8–10 and 14. Described data in the Results using Table 1 and also in the Methods. |
| 11. **Main results**   a) Report the associations between genetic variant and exposure, and between genetic variant and outcome, preferably on an interpretable scale (e.g. comparing 25th and 75th percentile of allele count or genetic risk score, if individual-level data available)  b) Report causal effect estimate between exposure and outcome, and the measures of uncertainty from the MR analysis. Use an intuitive scale, such as odds ratio, or relative risk, per standard deviation difference  c) If relevant, consider translating estimates of relative risk into absolute risk for a meaningful time-period  d) Consider any plots to visualize results (e.g. forest plot, scatterplot of associations between genetic variants and outcome versus between genetic variants and exposure) | Page 14–15. Reported in the “Severe COVID-19 outcome” section and the “COVID-19 hospitalization outcome” section. Also presented forest plots in Figure 3 and Figure 4. |
| 12. **Assessment of assumptions**   a) Assess the validity of the assumptions.  b) Report any additional statistics (e.g., assessments of heterogeneity, such as I2, Q statistic) | Page 16. Reported in the “Sensitivity analysis” section. |
| 13. **Sensitivity and additional analyses**   a) Use sensitivity analyses to assess the robustness of the main results to violations of the assumptions  b) Report results from other sensitivity analyses (e.g., replication study with different dataset, analyses of subgroups, validation of instrument(s), simulations, etc)  c) Report any assessment of direction of causality (e.g., bidirectional MR)  d) When relevant, report and compare with estimates from non-MR analyses  e) Consider any additional plots to visualize results (e.g., leave-one-out analyses) | Page 16. Reported in the “Sensitivity analysis” section. |
| **Discussion** |  |
| 14. **Key results**   Summarize key results with reference to study objectives | Page 16–17. Summarized key results in the 1^st^ paragraph of the Discussion. |
| 15. **Limitations**   Discuss limitations of the study, taking into account the validity of the MR assumptions, other sources of potential bias, and imprecision. Discuss both direction and magnitude of any potential bias, and any efforts to address them | Page 19–21. Discussed limitations in the 6^th^ paragraph of the Discussion section. |
| 16. **Interpretations**   a) Give a cautious overall interpretation of results considering objectives and limitations  Compare with results from other relevant studies.  b) Discuss underlying biological mechanisms that could be modelled by using the genetic  variants to assess the relationship between the exposure and the outcome  c) Discuss whether the results have clinical or policy relevance, and whether interventions  could have the same size effect | Page 17–19. Discussed the interpretation of the results in the 2^nd^–5^th^ paragraphs. |
| 17. **Generalizability:**   Discuss the generalisability of the study results (a) to other populations, (b) across other exposure periods/timings, and (c) across other levels of exposure | Page 21. Discussed the generalizability in the 6^th^ paragraph. |
| 18. **Funding:**   Describe sources of funding and the role of funders in the present study and, if applicable, sources of funding for the databases and original study or studies on which the present study is based | Page 21 and 22. Reported funding in the “Acknowledgments” section. |
| 19. **Data and data sharing:**   Provide the data used to perform all analyses or report where and how the data can be accessed, and reference these sources in the article. Provide the statistical code needed to reproduce the results in the article, or report whether the code is publicly accessible and if so, where | Page 22. Reported data sharing in the “Data availability” section. Provided genetic variants used as instrumental variables in the Supplementary table 1. |
| 20. **Conflicts of Interest:** All authors should declare all potential conflicts of interest | Page 3. Declared conflicts of interest in the “Disclosure summary” section. |
